# Meta-analysis and adjusted estimation of COVID-19 case fatality risk in India and its association with the underlying comorbidities

**DOI:** 10.1101/2020.10.08.20209163

**Authors:** Balbir B. Singh, Michael P Ward, Mark Lowerison, Ryan T. Lewinson, Isabelle A. Vallerand, Rob Deardon, Játinder PS. Gill, Baljit Singh, Herman W. Barkema

## Abstract

There is a lack of COVID-19 adjusted case fatality risk (aCFR) estimates and information on states with high aCFR. State-specific aCFRs were estimated, using 13-day lag for fatality. To estimate country-level aCFR, state estimates were meta-analysed. Multiple correspondence analyses (MCA), followed by univariable logistic regression, were conducted to understand the association between aCFR and geodemographic, health and social indicators. Based on health indicators, states likely to report a higher aCFR were identified. Using random- and fixed-effects models, the aCFRs in India were 1.42 (95% CI 1.19 – 1.70) and 2.97 (95% CI 2.94 – 3.00), respectively. The aCFR was grouped with the incidence of diabetes, hypertension, cardiovascular diseases and acute respiratory infections in the first and second dimensions of MCA. The current study demonstrated the value of using meta-analysis to estimate aCFR. To decrease COVID-19 associated fatalities, states estimated to have a high aCFR must take steps to reduce co-morbidities.

**Article Summary Line:** Meta-analysis and the COVID-19 adjusted case fatality risks (aCFRs) in India are reported and states likely to report a higher aCFR have been identified.

---

Novel severe acute respiratory syndrome coronavirus-2 (SARS-CoV-2) was first reported in Wuhan, China in December 2019 (1). Coronavirus disease 2019 (COVID-19) progressed rapidly into a serious pandemic and within 10 months, despite mitigation efforts, > 30M cumulative cases and 0.95M deaths due to COVID-19 have been reported worldwide (2). Furthermore, an unmitigated outbreak was estimated to cause ∼ 7B infections and ∼40 M deaths worldwide in 2020 (3).

SARS-CoV-2 is transmitted via respiratory droplets and aerosols from infected individuals (4, 5). Virus particles present in small droplets released while speaking or coughing can remain viable and infectious in aerosols for 3 hours (6, 7). These virus particles can be transmitted directly or by contact transfer via contaminated hands (8). Furthermore, transmission of SARS-CoV-2 has been linked to temperature and humidity (9, 10). COVID-19 symptoms include fever, cough, shortness of breath, pneumonia and other respiratory tract symptoms, and can progress to death (11, 12). The median incubation period is 5.1 days (95% CI, 4.5 to 5.8 days) and 97.5% will develop symptoms within 11.5 days (95% CI 8.2 – 15.6 days) of infection (13). Only ∼1% of cases will develop symptoms after 14 days of active monitoring or quarantine (13).

Case fatality risk (CFR) estimates for COVID-19 vary across countries and over time. As of 5 March 2020, a CFR of 3.5% was reported from China, 4.2% was reported across 82 countries/territories, and 0.6% from a cruise ship (after accounting for a lag time for fatality) (14). On 17 March 2020, a CFR of 7.2% was reported from Italy (15). A CFR of 5.65% (after accounting for right-censoring) was reported in mainland China, based on data collected from 29 December 2019 to 17 April 2020 (16). Case fatality rates of 1.2% in Germany (17), 9% in Spain, 11.9% in Italy, 8.6% in the Netherlands, 7.1% in France and 8% in the UK, have been reported across varying intervals (18). Thus, there appears to be great variability among CFR estimates, from < 1 to > 10%.

COVID-19 prognosis and progression vary among individuals. Diabetes has been reported to be a risk factor for a poor COVID-19 prognosis (19). Coronary artery disease, heart failure, cardiac arrhythmia, chronic obstructive pulmonary disease, current smoking and > 65 years of age were associated with an increased risk of in-hospital death among COVID-19 hospitalized patients (20). A higher frequency of obesity was reported in intensive care COVID-19 patients (21).

The first case of COVID-19 in India was reported on 30 January 2020 (22). As of 22 September 2020, the country had reported 5,562,663 cumulative cases, a total of 88,935 deaths and >65M tests conducted (23). The disease has been reported in all states and union-territories. To our knowledge, state-specific CFRs (after accounting for a lag-time for fatality) and the case fatality risk at the country level using a meta-analysis approach have not been reported. In addition, little is known about the risk factors associated with COVID-19 CFR in India. Therefore, our objectives were to estimate the aCFR for COVID-19 and its association with various health and social indicators in India.

## Methods

### Source of data

#### COVID-19 case and death data

State/Union territory-specific COVID-19 cumulative case (27 July 2020 – 08:00 AM IST) and death (27 July 2020 – 08:00 AM IST and 09 August 2020 – 08:00 AM IST) time-series data were extracted from the Ministry of Health and Family Welfare and Indian Council for Medical Research, Government of India websites (23).

Combined data for the union territories Jammu and Kashmir and Ladakh are presented (old state of Jammu and Kashmir), as other health and social indicators were only available for the old state of Jammu and Kashmir. No data were available for UT of Lakshadweep. Furthermore, individual data for the two UTs Dadra and Nagar Haveli and Daman and Diu were not available and were therefore excluded. Mizoram state had limited cases with no deaths reported; therefore, this state was also excluded from the analysis. The final dataset had information on 32 states/union territories (Fig. 1).

**Fig. 1.**
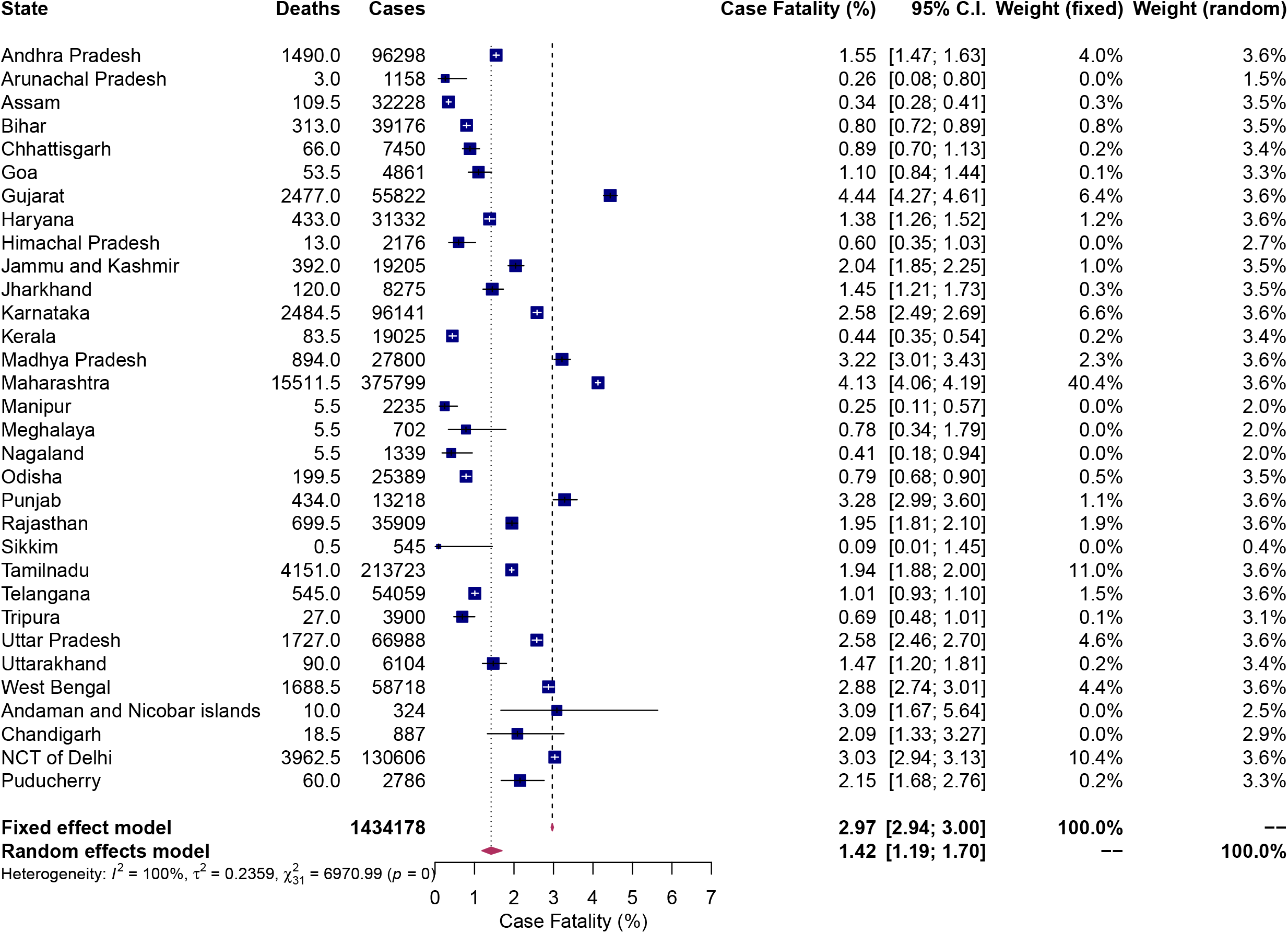
Forest plot of case fatality risk of COVID-19 in India, using random- and fixed-effect models.

#### State/UT-specific parameters

Data for 17 state-specific geodemographic (n=4), socio-economic (n=1), health and comorbidity-related (n=12) factors were collected (Table 1). Information on the 2016 projected total human population and number of persons who attended Non-communicable disease (NCD) clinics in 2018 were also collected to derive estimates per 10,000 population (Table 1).

**Table 1.**
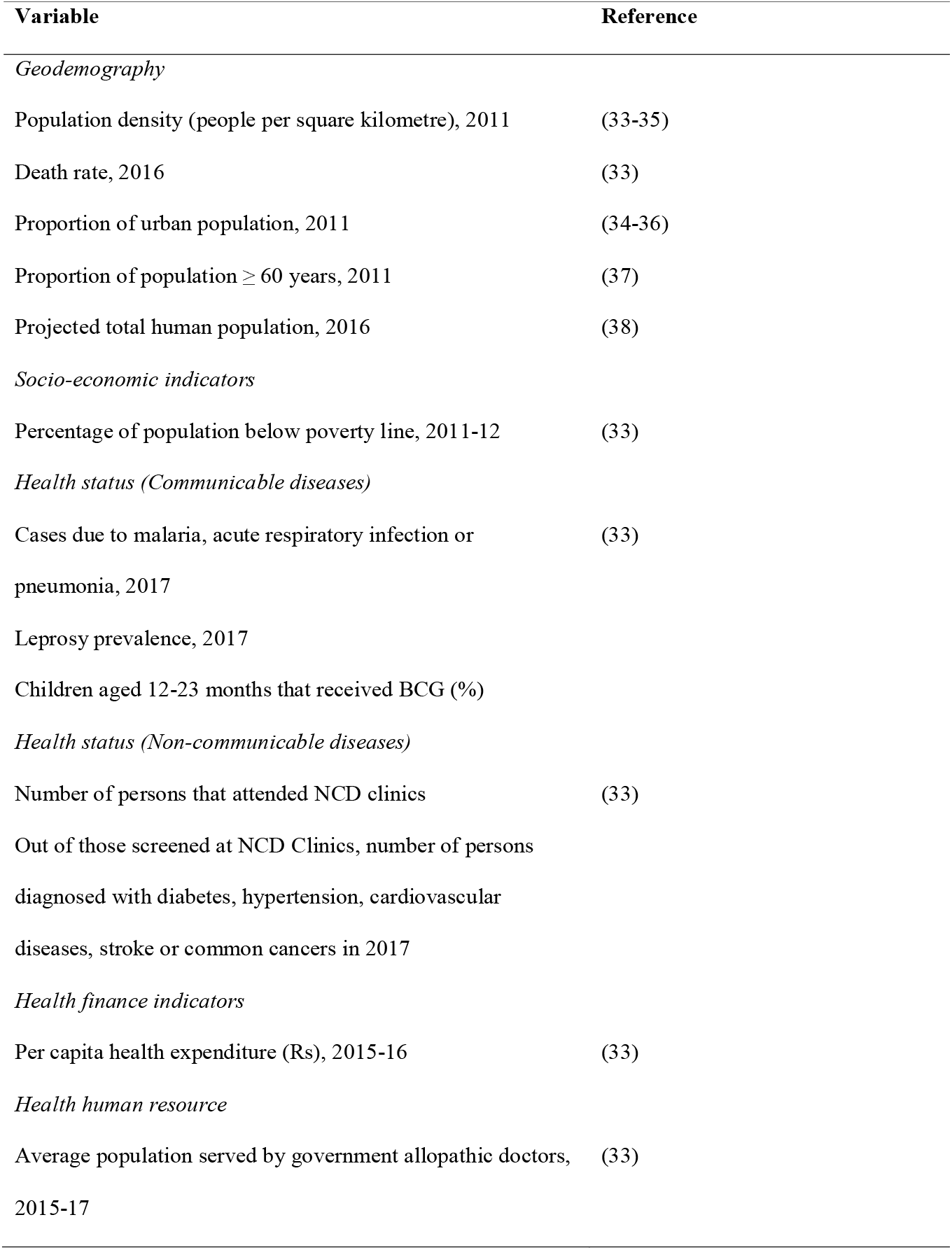
Country-specific geodemographic, environment, social, health and comorbidity-related variables used in the study.

### Database development

Annual incidence of malaria, acute respiratory infection, and pneumonia per 10,000 human population were estimated by dividing the total number of cases reported due to malaria, acute respiratory infection, and pneumonia in 2017 by the projected human population in 2016, and finally multiplied by 10,000.

Similarly, incidence of diabetes, hypertension, cardiovascular diseases, stroke, and common cancers (from those who attended NCD clinics) were divided by the total number of persons attending NCD clinics and finally multiplied by 10,000 to estimate number of patients diagnosed with diabetes, hypertension, cardiovascular diseases, stroke, and common cancers per 10,000 population that visited NCD clinics.

### Statistical analyses

Statistical analyses were conducted using R (R statistical package version 3.4.0, R Development Core Team [2015], http://www.r-project.org).

#### Adjusted case fatality risk (aCFR)

State-specific aCFR was estimated using a lag time for fatality. We used a previously reported median time delay of 13 days from illness onset to death (24). The number of COVID-19 cumulative cases on 27 July 2020 was used as the denominator value. Based on Wilson, Kvalsvig (14), we assumed that half of the additional cumulative reported deaths on 9 August 2020 corresponded to cases reported on 27 July. As noted, this approach is simple, albeit likely to be modified or replaced accurate studies to overcome associated limitations become available (14).

Finally, the aCFR (proportion of cumulative deaths to cumulative cases) was estimated via meta-analysis using the R function *metaprop*, with inverse-variance weighting. Separate estimates using both fixed- and random-effects models are presented (25). Proportions were logit transformed.

#### Predictors and outcome

Information on the 17 state-specific geodemographic, social, health and comorbidity-related factors as key predictors were collected. State-specific aCFR was used as the outcome variable.

#### Descriptive analyses

Descriptive analyses were performed and data were tested for the assumptions of linearity and normality. Initially, a variable was log-transformed if the assumption of normality was not met. Later, non-linear variables were converted into categorical variables using quartiles for further analysis. Fisher’s Exact test was used to determine the association between categorical predictor variables.

#### Multiple correspondence analyses

Due to expected associations among categorical predictor variables, multiple correspondence analysis (MCA) was conducted to determine potential groupings of predictor variables with the aCFR. For the purpose of MCA, categorical predictor variables were re-categorised using their medians and converted into dichotomous low- or high-value predictor variables.

Adjusted CFR was also dichotomised as low versus high aCFR before conducting the MCA. States with missing values were excluded from the MCA.

#### Univariable logistic regression

Predictor variables grouped with aCFR (in the same quadrant as the group centroid in the first- and second dimensions following MCA) were assessed using univariable linear regression (p ≤ 0.05). In addition, incidence of patients with stroke that visited NCD clinics was also assessed as it was placed closer to the variable aCFR in the MCA (albeit in a different quadrant). Dichotomised aCFR was used as the outcome variable and the selected geodemographic and health indicators (after categorising by their quartiles) as key predictors.

#### Identification of states with high aCFR

Based on the subjective evaluation of the univariable analysis (using significant predictor variables), states having a low, medium, or high aCFR were determined. The predictor quartile having the lowest odds ratio was assigned a score of 1 and that of the highest odds ratio was assigned a score of 4. Predictor quartiles having similar odds ratios were assigned average scores for their respective rank quartiles. Scores of all the significant predictors were combined to produce an overall risk score of aCFR. States having scores of 0–8, 8–16 and 16–24 were categorised as low, medium, and high risk aCFR states. Choropleth maps describing risk score of aCFR for the Indian states were generated.

## Results

### Case fatality risk

Overall, in the selected states/union territories by 27 July 2020 1,434,178 cumulative COVID-19 cases had been reported, whereas by 9 August 2020 43,377 deaths had been reported. Using the random-effects model and meta-analysis, the aCFR (%) was estimated to be 1.42 (95% CI 1.19 – 1.70). Furthermore, using the fixed-effects model and meta-analysis, the aCFR was estimated to be 2.97 (95% CI 2.94 – 3.00). Heterogeneity was very high at 99.57% (p < 0.001) in the effect of sizes in both fixed- and random-effect models (Fig. 1). Based on high heterogeneity, random-effects model estimates were more likely to be representative of the true aCFR for India.

### Multivariable correspondence analysis

Case fatality risk grouped with certain geodemographic and health indicators in the first and second dimensions of MCA (Fig. 2); potentially associated variables listed (Table 2). Grouping of aCFR with the incidence of diabetes, hypertension, cardiovascular diseases and acute respiratory infections was apparent in the first and second dimensions of the multiple correspondence analysis.

**Table 2.**
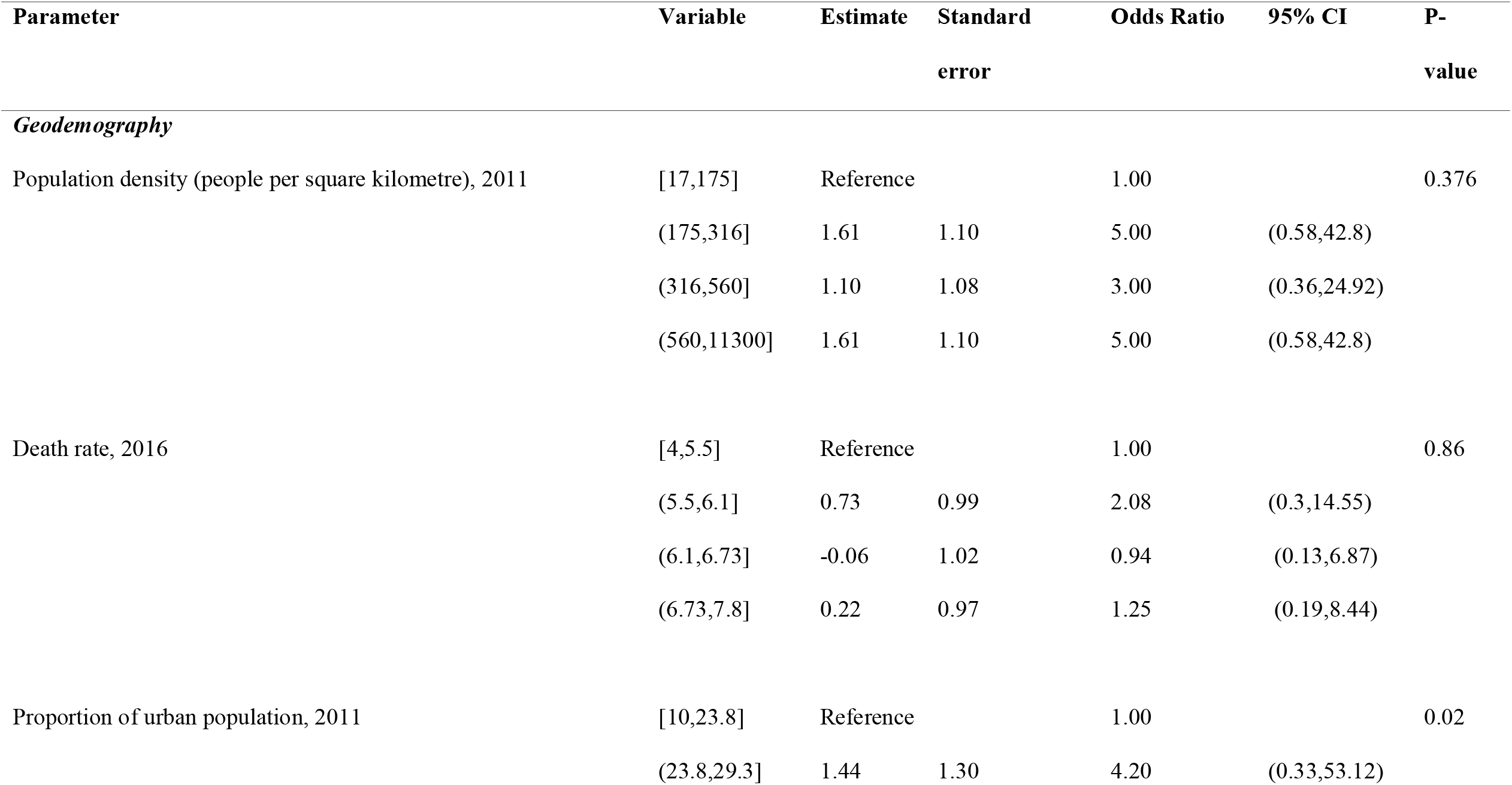
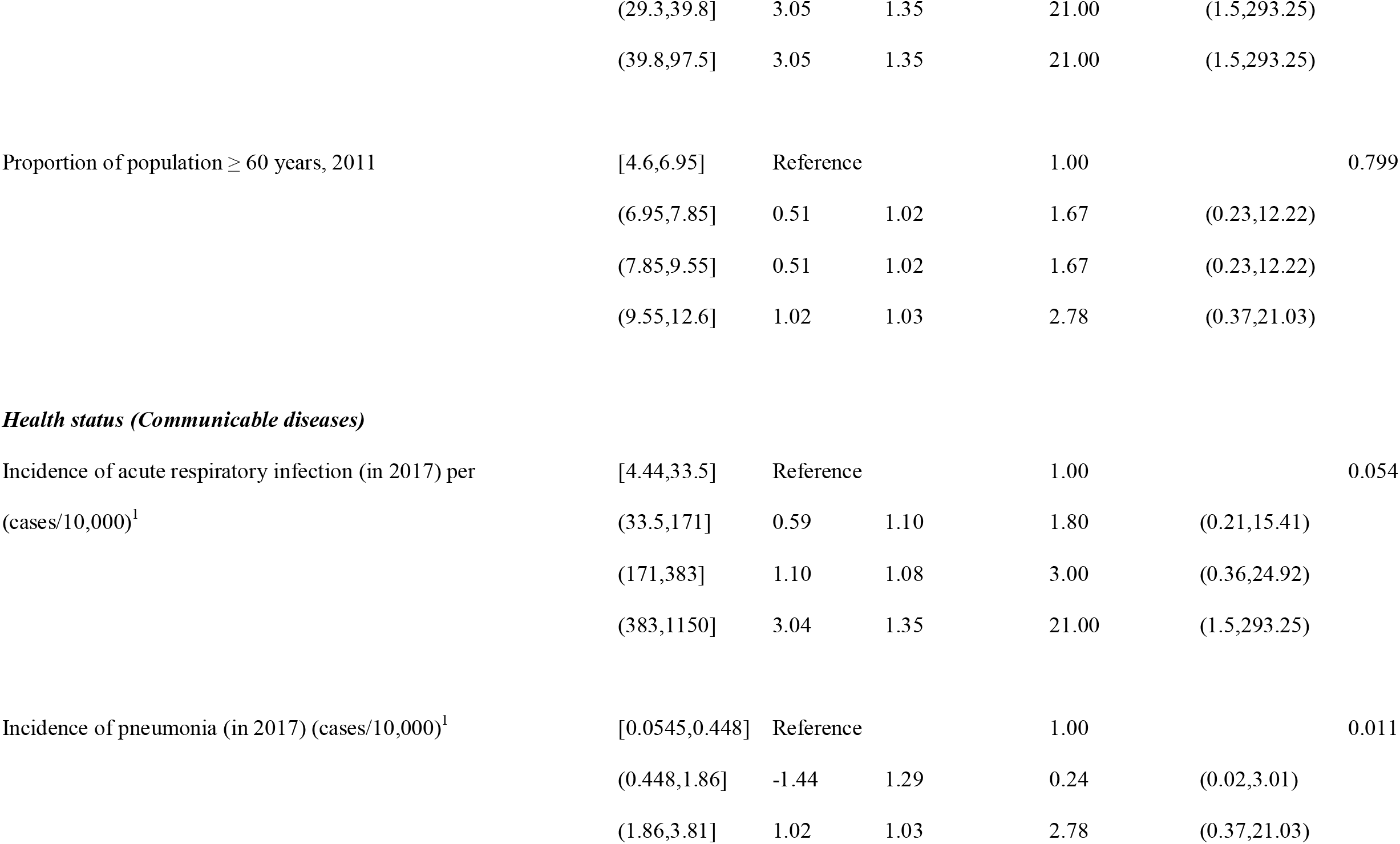
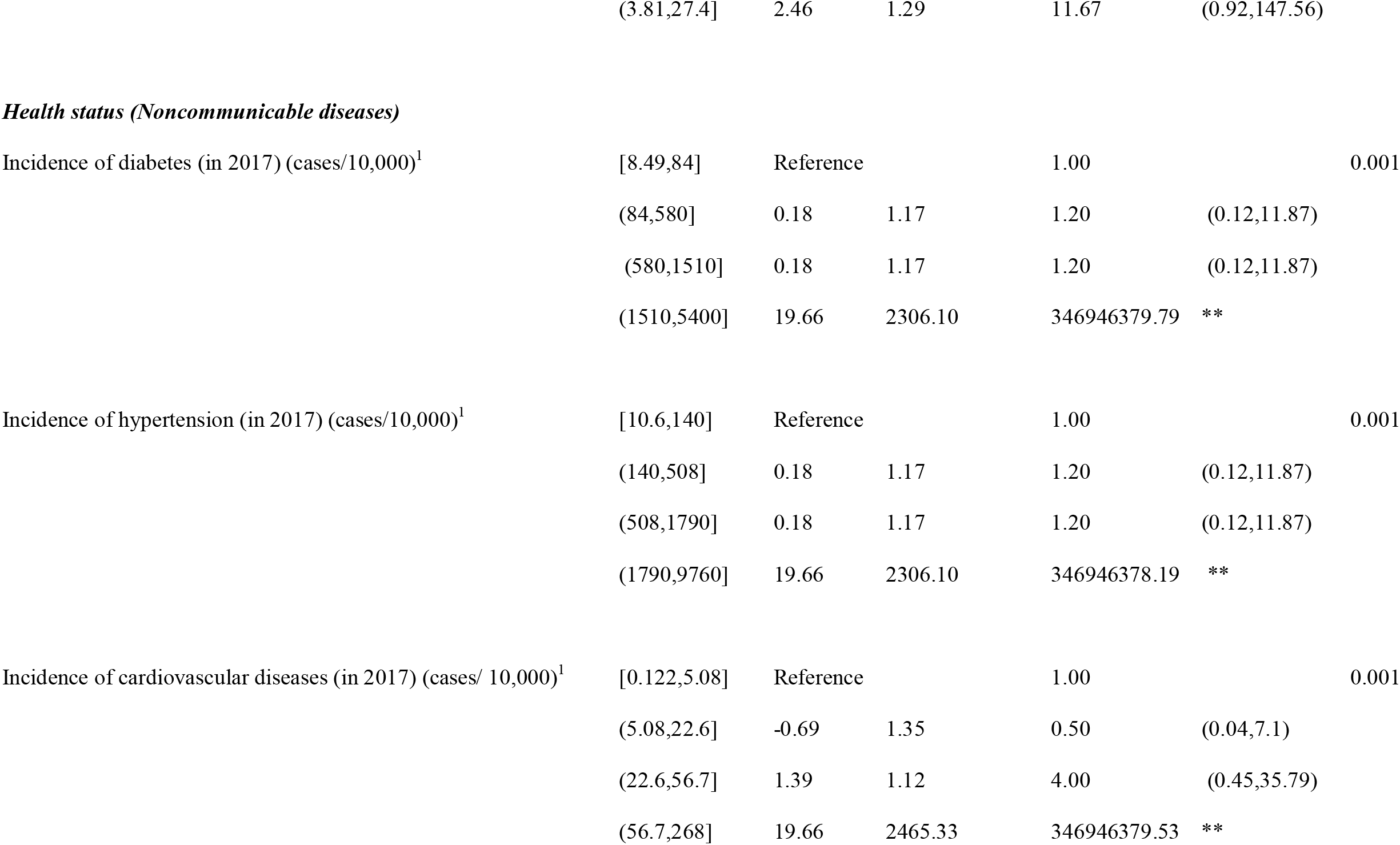
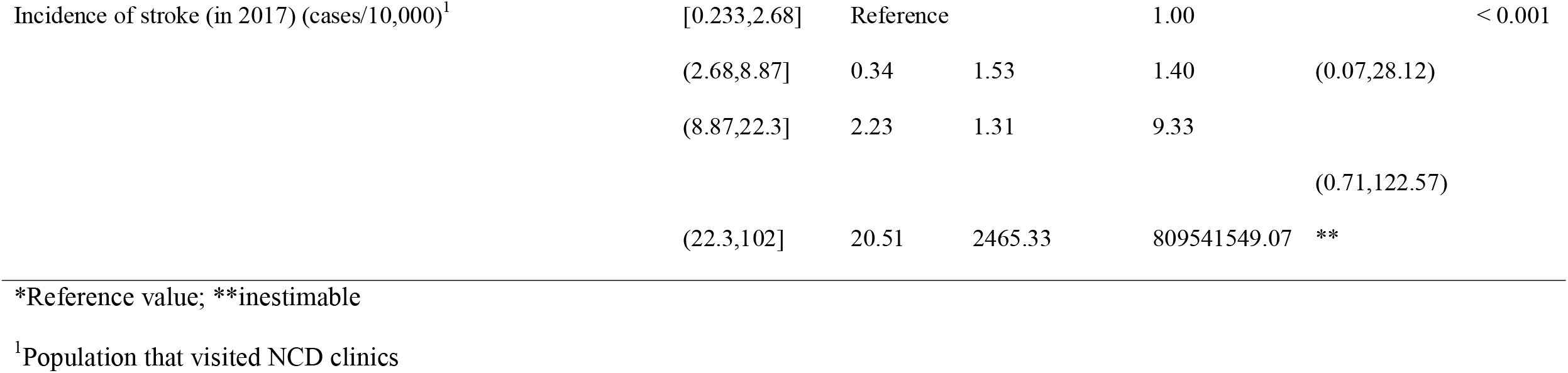
Summary of univariable logistic regression analyses of state-specific geodemography and health indicators associated with adjusted COVID-19 case fatality risk in India

**Fig. 2.**
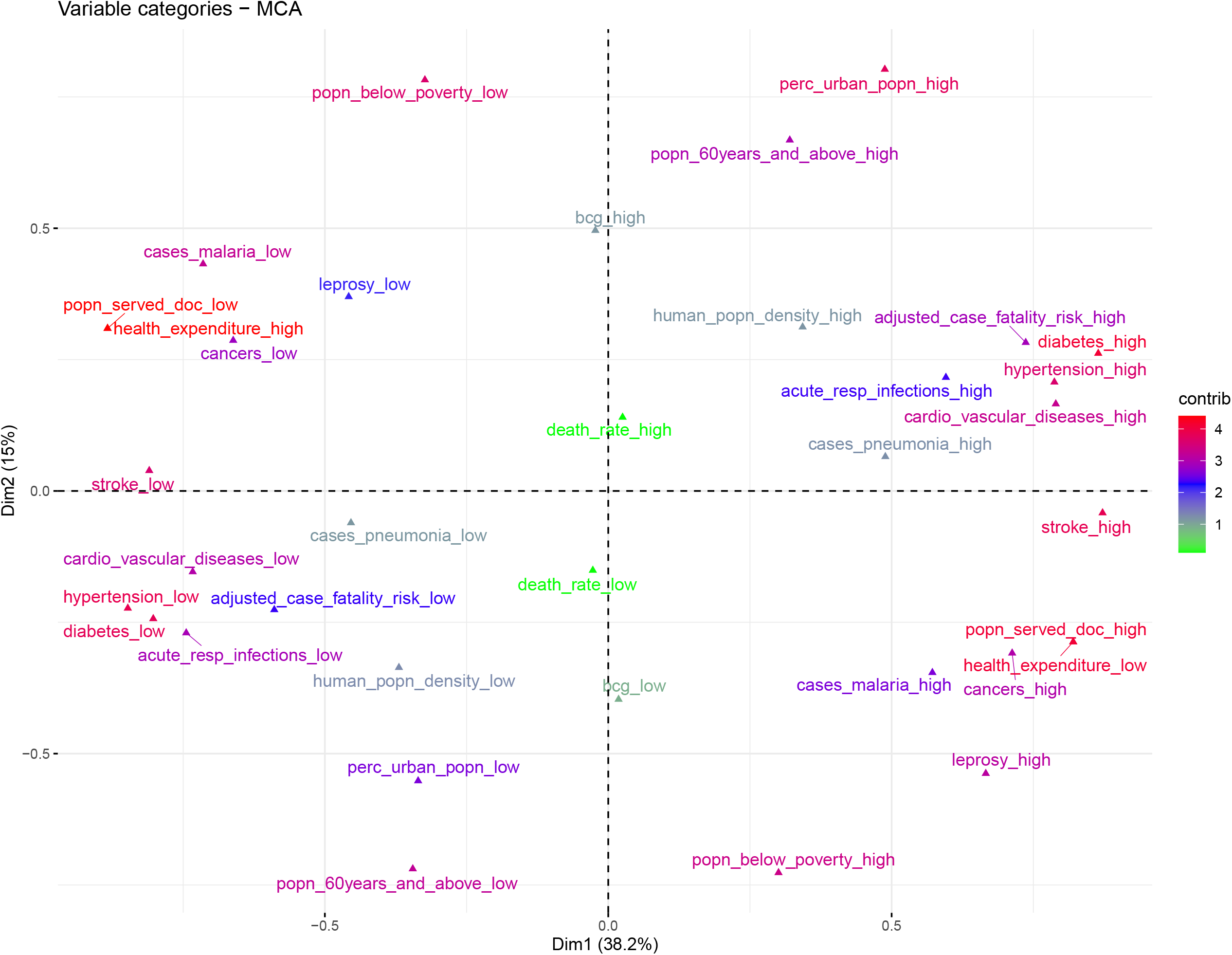
Plot of the first and second dimensions of multiple correspondence analysis of state-specific case fatality risk and geodemography, social and health indicators in India.

### Univariable analysis

Univariable analysis revealed that the 2011 proportion of urban population, 2017 incidence of diabetes, 2017 incidence of hypertension, 2017 incidence of cardiovascular diseases, 2017 incidence of stroke, and 2017 incidence of pneumonia were positively correlated with COVID-19 aCFR (Table 2).

### Identification of states with high aCFR

Based on the predictor variables, we categorised 10, 17 and 1 Indian states as likely to have a high, medium, and low aCFR risk, respectively (Fig. 3, Table 3).

**Table 3.** Qualitative risk evaluation of the adjusted case fatality risk (aCFR, %) in different states of India.

| State | Proportion of<br>urban population,<br>2011 | Incidence of specific diseases in 2017 |  |  |  |  | aCFR<br>(%) | Risk<br>score-<br>aCFR | Risk score-<br>aCFR level |
| --- | --- | --- | --- | --- | --- | --- | --- | --- | --- |
|  |  | Pneumonia | Diabetes<sup>1</sup> | Hypertension<sup>1</sup> | Cardiovascular<br>diseases<sup>1</sup> | Stroke<sup>1</sup> |  |  |  |
| Andhra Pradesh | 3.5 | 4 | 4 | 4 | 4 | 4 | 1.55 | 23.5 | High |
| Arunachal Pradesh | 1 | 2 | 1 | 2.5 | 2 | 2 | 0.26 | 10.5 | Medium |
| Assam | 1 | 3 | 2.5 | 2.5 | 3 | 3 | 0.34 | 15 | Medium |
| Bihar | 1 | 4 | 2.5 | 1 | 1 | 1 | 0.80 | 10.5 | Medium |
| Chhattisgarh | 1 | 1 | 2.5 | 2.5 | 1 | 2 | 0.89 | 10 | Medium |
| Goa | 3.5 | 1 | 2.5 | 2.5 | 1 | 2 | 1.10 | 12.5 | Medium |
| Gujarat | 3.5 | 2 | 4 | 4 | 4 | 4 | 4.44 | 21.5 | High |
| Haryana | 3.5 | 1 | 2.5 | 2.5 | 1 | 2 | 1.38 | 12.5 | Medium |
| Himachal Pradesh | 1 | 3 | 2.5 | 2.5 | 1 | 1 | 0.60 | 11 | Medium |
| Jammu and Kashmir | 2 | 3 | 2.5 | 2.5 | 1 | 2 | 2.04 | 13 | Medium |
| Jharkhand | 2 | 1 | 2.5 | 2.5 | 3 | 3 | 1.45 | 14 | Medium |
| Karnataka | 3.5 | 3 | 2.5 | 2.5 | 3 | 3 | 2.58 | 17.5 | High |
| Kerala | 3.5 | 1 | 2.5 | 2.5 | 3 | 2 | 0.44 | 14.5 | Medium |
| Madhya Pradesh | 2 | 4 | 2.5 | 2.5 | 4 | 3 | 3.22 | 18 | High |
| Maharashtra | 3.5 | 1 | 4 | 4 | 3 | 4 | 4.13 | 19.5 | High |
| Manipur | 2 | 2 | 1 | 1 | 2 | 1 | 0.25 | 9 | Medium |
| Meghalaya | 1 | 1 | 1 | 1 | 2 | 1 | 0.78 | 7 | Low |
| Nagaland | 2 | 2 | 1 | 1 | 2 | 1 | 0.41 | 9 | Medium |
| Odisha | 1 | 3 | 2.5 | 2.5 | 1 | 3 | 0.79 | 13 | Medium |
| Punjab | 3.5 | 3 | 4 | 4 | 4 | 4 | 3.28 | 22.5 | High |
| Rajasthan | 2 | 4 | 4 | 4 | 4 | 4 | 1.95 | 22 | High |
| Sikkim | 2 | 2 | 1 | 1 | 2 | 1 | 0.09 | 9 | Medium |
| Tamil Nadu | 3.5 | 3 | 4 | 4 | 3 | 4 | 1.94 | 21.5 | High |
| Telangana | 3.5 | 1 | 2.5 | 2.5 |  |  | 1.01 | NA | NA |
| Tripura | 2 | 2 | 1 | 1 | 2 | 1 | 0.69 | 9 | Medium |
| Uttar Pradesh | 1 | 4 | 4 | 4 | 4 | 3 | 2.58 | 20 | High |
| Uttarakhand | 3.5 | 3 | 2.5 | 2.5 | 2 | 1 | 1.47 | 14.5 | Medium |
| West Bengal | 3.5 | 4 | 4 | 4 | 4 | 4 | 2.88 | 23.5 | High |
| Andaman and<br>Nicobar<br>Islands | 3.5 | 2 |  |  |  |  | 3.09 | NA | NA |
| Chandigarh | 3.5 | 4 | 1 | 1 | 2 |  | 2.09 | NA | NA |
| NCT of Delhi | 3.5 | 4 |  |  |  |  | 3.03 | NA | NA |
| Puducherry | 3.5 | 2 | 1 | 1 | 3 | 3 | 2.15 | 13.5 | Medium |

Ranks: Low (0-8), Medium (8-16), High (16-24), NA – No rank assigned
<sup>1</sup>Cases/10,000 that visited NCD clinics.

**Fig. 3.**
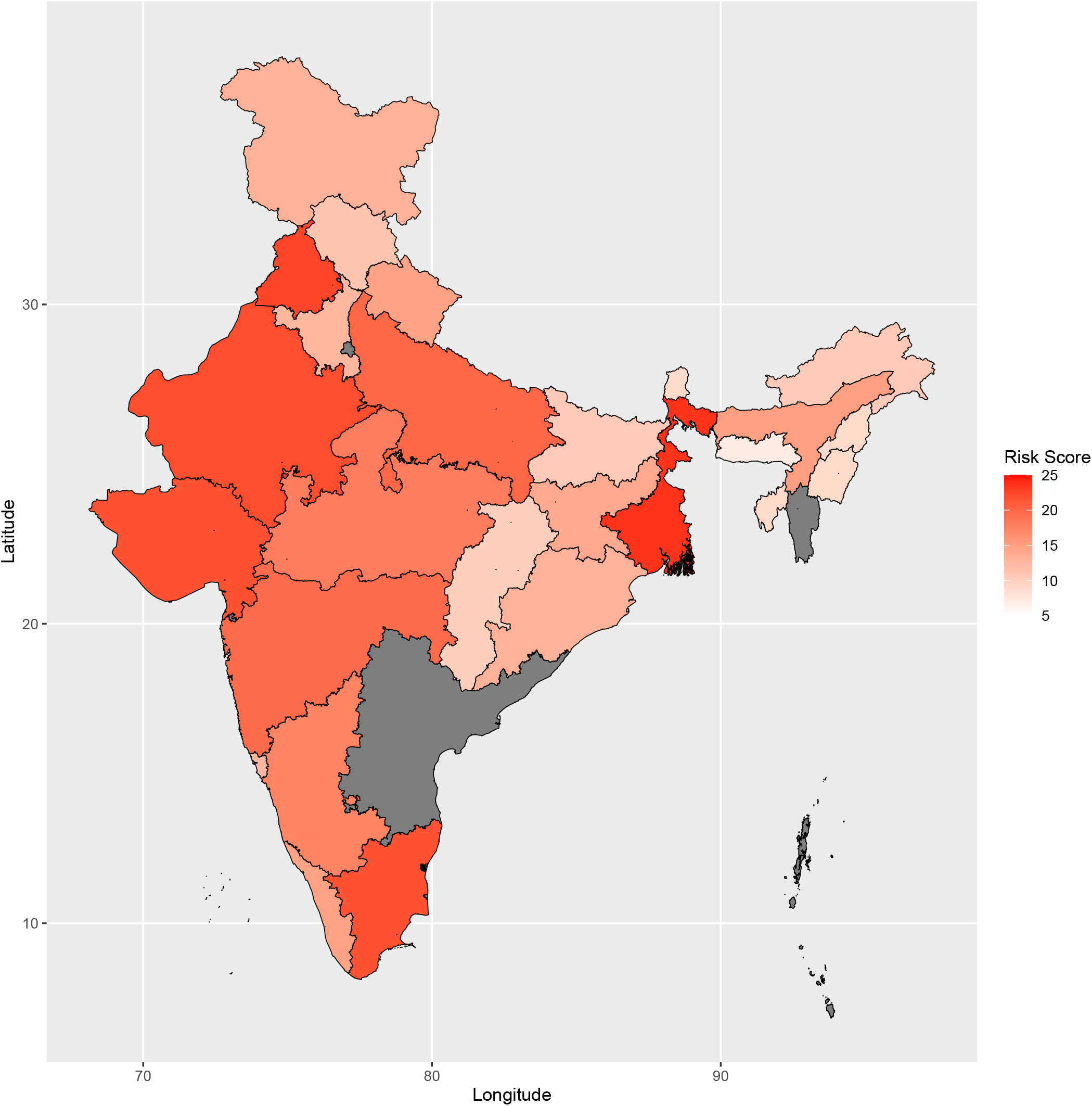
Estimated risk score of the adjusted case fatality risk (aCFR) for various states of India.

## Discussion

To our knowledge, the aCFR, using meta-analysis and a lag time for fatality using state-specific data, has not been estimated for India or many other countries. However, this will help inform COVID-19 response in the country. Similarly, identifying states with high CFR will help to better allocate health resources across states and enhance preparedness levels in these states.

Accurate estimation of CFR is a serious challenge worldwide. We used a previously reported median time delay of 13 days from illness onset to death and accounted for half of the deaths during this interval in aCFR estimations. Any bias in this estimate may have under- or over-estimated the COVID-19 aCFR in India. However, we believe this estimate to be much more accurate than the crude CFR estimations using same day COVID-19 case and death data. We agreed with a previous study that this approach is simple, albeit likely to be superseded when accurate studies to overcome associated limitations become available (14). In addition, asymptomatic cases, testing criteria and capacity further complicate COVID-19 case estimations.

Using the random- and fixed-effect models, the estimated aCFR was 1.42 (95% CI 1.19 – 1.70) and 2.97 (95% CI 2.94 – 3.00), respectively. Due to high heterogeneity, estimates using the random-effects model were more likely to represent the true aCFR for India. Previous studies used random-effect models to estimate the CFR of COVID-19 (26, 27) or presented CFR using both random-effects and fixed-effect models (28). Using a random effects model, we ensured that states with high numbers of cases and deaths received more weight compared to states with fewer cases and deaths.

For India, the aCFR appeared to be lower than in many European countries. This might be due to the fact that only 6.38% of the population in India was above 65 years of age in 2019 (29). Elderly people (>60 years) have been reported to be at a higher risk of death due to COVID-19 (30, 31). However, many other health and social indicators, changes in the virulence of SARS-CoV2 in regions over time, and country-specific COVID-19 response indicators may be associated with CFR, and this needs to be investigated.

Heterogeneity was as high as 99.57% in the effect sizes in state-specific aCFRs (both fixed- and random-effect models), perhaps due to differences in state-specific sub-populations, health facilities, infrastructure hospital care and COVID-19 testing protocols. In addition, specific states might be in different phases of the COVID-19 epidemic, as the CFR has varied during the progression of the COVID-19 epidemic (28). High heterogeneity needs to be further investigated; regardless, the current estimates suggest uncertainty of aCFR point estimates.

Multiple correspondence analyses and univariable logistic regression were used to summarise associations between state-specific aCFR and several geodemography, social and health indicators. Based on these analyses, health indicators such as incidence of diabetes, hypertension, stroke and cardiovascular diseases were associated with the aCFR in India, consistent with previous studies in other countries (19, 20).

The current study had some limitations. Differences in COVID-19 testing and hospital care may have caused differences in aCFR at the state level. Although health indicators were estimated from NCD clinic data, there may be differences among states in the population that visit NCD clinics. The Government of India has NCD clinics for screening and early diagnosis of NCDs under the National Programme for Prevention and Control of Cancer, Diabetes, Cardiovascular Diseases and Stroke. As of March 2020, 665 District NCD Cells, 637 District NCD Clinics, 4472 CHC NCD Clinics, 181 Cardiac Care Units and 218 Day Care Units were reported to be functional (32). Overall, 35.72M patients have been reported to attend NCD clinics in 2017 (33). In addition, the role of many health indicators, e.g. obesity, could not be evaluated. Furthermore, we could not account for the migration between the states. That COVID-19 testing capacity varies across states may have influenced the aCFR estimations in the current study. Lastly, the risk factor investigation at the state level may not be applicable at the individual level. Despite these shortcomings, we believe the aCFR and the risk factor investigation in the current study to be sufficiently valid to inform COVID-19 response in India and populations in similar settings.

## Data Availability

The corresponding author had full access to all data in the study and will be supplied on request by the corresponding author.

## Acknowledgements

The authors acknowledge India’s National and State Health departments for collecting the daily COVID-19 epidemic data and releasing it in the public domain.

The authors declare no conflicts of interest.

## Author Bio

Balbir Bagicha Singh is an Associate Professor at the Guru Angad Dev Veterinary & Animal Sciences University (India), Honorary Lecturer at the Sydney School of Veterinary Science (The UOS, Australia), and Adjunct Professor at the University of Saskatchewan (Canada). His primary research interest is the epidemiology of zoonotic and emerging infectious diseases.

